# Acceptability of Community Midwives in rural communities of Pakistan: A qualitative exploratory study

**DOI:** 10.1101/2023.02.02.23285378

**Authors:** Bakhtawar Muhammad Hanif Khowaja, Anam Shahil Feroz, Musarrat Rani, Quratulain Khowaja, Mehmooda Afroz Ali Khowaja, Farina Abrejo, Sarah Saleem

## Abstract

**Introduction:** The Government of Pakistan initiated the Community Midwifery program in 2006 to provide skilled birth attendance to women living in rural areas. However, the acceptability of midwives and what impact these community midwives have made on overall maternal morbidity and mortality remains an unanswered question. We explored the perceptions of health officials, midwifery students, midwives and community women about the factors that influence the acceptability of community midwives’ services in the rural district Thatta, Pakistan.

**Materials and Methods:** A qualitative exploratory study was conducted in the rural district Thatta of Pakistan. In-depth interviews were conducted with health officials, midwifery students who were currently enrolled in the midwifery program of the district; community midwives providing services in district Thatta, and trained community midwives who are not practicing. Interviews were also conducted with community women. Data were analyzed using the qualitative thematic analysis approach and the deductive analysis method.

**Results:** Two overarching themes were identified: (I) community acceptance and support; and (II) dynamics between CMWs and other health care providers. The major hindering factors to CMWs acceptance included their young age challenging social acceptability, patronizing behavior of doctors, high acceptance of traditional birth attendants working in rural areas, and the community’s reluctance towards referral services. The facilitating factors included clients’ privacy maintained at birth stations and the affordability of community midwives’ services.

**Conclusion:** There are deep-rooted challenges related to the acceptability of midwifery services at the community level and with other competing healthcare providers which need advocacy to support and accept their services at the community level and by other professionals.

## Introduction

It is estimated that globally around 830 women die every day due to preventable causes related to pregnancy and childbirth [1]. Around 73 low-to middle-income countries (LMICs) account for 92% of maternal and newborn mortalities including India and Pakistan [2]. Pakistan is among the top ten countries contributing 59% of maternal deaths globally [3]. The lifetime risk of death for a Pakistani woman due to pregnancy-related causes is 1 in 80, compared to 1 in 61 in low to middle-income countries including Africa and South Asia countries, and 1 in 4,085 in high-income countries [4].

Reduction in maternal mortality is a priority target under goal 3 of the Sustainable Development Goals (SDGs) i.e. “To ensure healthy lives and promote well-being for all at all ages” through 2030 [5]. Skilled birth attendance is a prerequisite to attaining a reduction in maternal mortality [6]. World Health Organization (WHO) emphasizes skilled care at birth to reduce maternal and newborn deaths [6]. According to the WHO, skilled health workers include a trained doctor, nurse, or midwife during pregnancy, delivery, and postnatal period [7].

There is collective evidence about the role of midwives in positive pregnancy outcomes for women and newborns [8]. Accessible midwifery services reduce health inequalities across the world by provision of services to underserved areas and it offers positive benefits in terms of economic, efficient, and effective services [9]. Most countries succeeded in improving maternal mortality and morbidity by ensuring the availability and accessibility of midwifery services and primary care thus improving skilled care for pregnant women [10].

Sweden midwives are well recognized and have unique history regarding the involvement of midwives in their health system. The intense reduction in maternal mortality in Sweden during the last century has been attributed to the role of midwifery [10]. This has been achieved through midwives’ contribution to all aspects of women’s health care [10]. Swedish midwives hold a unique level of autonomy in the health system to provide services during pregnancy and childbirth [10].

Based on the evidence of community midwives’ contribution to addressing the issue of high maternal mortality, many countries adopted the model of introducing community midwives into their health systems [11]. The Government of Pakistan also initiated the Community Midwifery program in 2006 to provide skilled birth attendance to women living in rural areas [12]. The CMW program was introduced to establish home-based clinics to provide antenatal, postnatal, and neonatal services to rural communities [12].

Despite a large investment in the CMW program by the government of Pakistan, the challenge of the non-availability and non-accessibility of community midwives in their local communities is apparent. Research evidence from rural districts of Pakistan suggests that the utilization of maternal and newborn services through community midwives is very low and that a large proportion of trained CMWs are not in practice. [13].

Reports from 2012 to 2013 show that only 20.8 % of pregnant women in district Thatta received maternal healthcare services [19]. Thus, the midwifery school was established in the year 2013 in the district to train community midwives for the provision of maternal and neonatal services [20]. The midwifery school offers 24 months diploma program and trains approximately more than 30 CMWs each year [20]. However, the district data displays that out of 150 deployed and registered CMWs, only 17 CMWs report to the maternal and neonatal, child heath (MNCH) program monthly. Identifying the causes of CMWs’ non-retention and non-availability in their communities is important to inform suitable policy recommendations.

To do so, we focused on the views of health officials, community midwives, and community women of district Thatta to explore factors influencing community midwives’ utilization of services through the use of the Community Midwifery (CMW) model [16]. The CMW model (Figure. 1) has been established with the Government of Pakistan’s MNCH program strategy to expand community-level CMWs services in Pakistan [15].

**Fig. 1.**
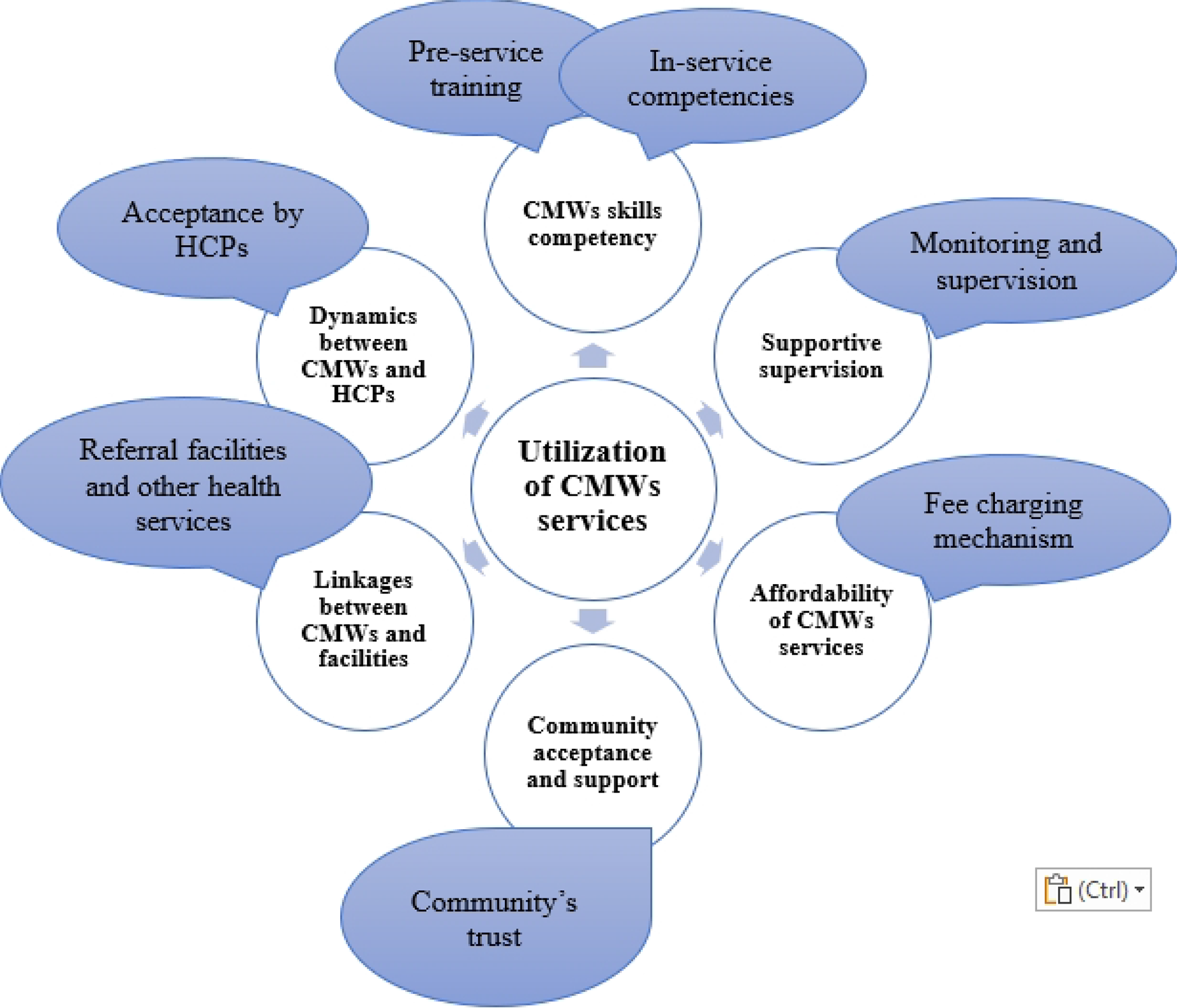
Community Midwifery Model

It comprises six major factors: (I) CMWs skills competencies, (II) supportive supervision, (III) affordability for CMWs services, (IV) community acceptance and support, (V) linkages between CMWs and facilities, and (VI) dynamics between CMWs and health care providers.

The findings on facilitators and barriers to the factors of supportive supervision and CMWs’ skills and competencies have been reported in a separate paper [17]. In this paper, we present results on the factors related to CMWs acceptability to practice in their communities.

## Materials and Methods

### Study setting

The study site selected was district Thatta of Province Sindh, which is a rural district with a total population of 979,817 [18]. The number of community health centers available in the district is adequate [25] and there is a midwifery school to train midwives [26]. The number of trained community midwives is sufficient in the district as evident by the health profile of the district [27], however, the services provided by community midwives is unknown.

### Study design and participants

We used an exploratory qualitative study design by conducting in-depth interviews (IDIs) using a semi-structured interview guide and a purposive sampling approach. The study is stated as per the guidance provided in the consolidated criteria for reporting qualitative research (supplemental file 1).

IDIs were conducted with officials from the health department who were involved in CMWs training and deployment including the district health officer, the district coordinator of the MNCH program, the principal of the midwifery school of district Thatta, the general secretary of the midwifery association of Pakistan, and the director of general nursing and midwifery program. The aim of interviews with health officials and program implementers was to understand their roles related to administrative and organizational perspectives.

Interviews were also conducted with three categories of the CMWs including midwifery students who were currently enrolled in the community midwifery program in Thatta; trained community midwives providing services in district Thatta; trained community midwives who have left the midwifery profession; and with community married women from district Thatta. The purpose of interviews with health officials, students, midwives, and community women was to explore key factors encountered at each level of midwifery training and practice.

### Data collection and reflexivity

Semi-structured interview guides were developed by the research team and reflected the four domains of the CMW model on the acceptability of CMWs. The detailed interview guides are provided as Supplemental file 2. These were prepared in the English language and translated into local languages Urdu and Sindhi to capture the perceptions of the study participants. The interviews were conducted in Urdu and Sindhi languages between August 2020 and December 2020 by the investigating team. All the study team members have expertise in qualitative research and an understanding of the scope of practices of midwives for maternal and child health.

We included key themes with several specific questions: (1) Affordability for CMWs services, such as CMWs fee for services, financial class of the community that approaches CMWs, community women affordability to utilize services (2) Community’s acceptance and support, such as the community’s trust for CMWs services, women preference for maternal and neonatal services (3) Linkages between CMWs and facilities, such as linkages with referral health facilities (4) Dynamics between CMWs and other health care providers, such as their relationships with doctors, lady health workers, traditional birth attendants, sonologists, dispensers, and other healthcare workers working in the region.

Due to the restrictions in the district because of the Covid-19 situation, all the interviews were conducted via telephonic calls. These interviews were scheduled according to participants’ preference of time and were audio-recorded following verbal informed consent from study participants. The purpose of the study was well-informed to all participants through an official letter sent by the university in advance of their interviews. Each interview lasted between 45 to 60 minutes. The lead author was fluent in local languages and carried out all the interviews. The data collection was carried out until data saturation was achieved and no new information emerged. We defined saturation as the amount of data needed until no new information and meaningful conclusions were drawn out on the research questions [22].

### Data analysis

The data were transcribed into Sindhi and Urdu languages and later translated into English language and analyzed manually. The qualitative thematic analysis approach and the deductive analysis method were adopted as an ongoing iterative process through which facilitating and hindering factors regarding CMWs’ acceptability were identified. This allowed for rigorous probing during interviews to collect the maximum data about the topic.

Manual analysis was done with familiarization of the data which involved reading and re-reading the transcripts to get familiar with the content. Codes were generated to identify important features of the data that were relevant to answering the research questions. The entire dataset was coded, and all the codes and relevant data extracts were collated. The codes and the collated data were examined for each potential theme. The themes were reviewed in relation to the coded extracts and the entire data set to generate a thematic map.

The researchers from the investigating team (SS, AF, FA, BK,) participated in refining the codebook to reach a final consensus and resolve any discrepancies to improve the credibility and reliability of the data [23]. To adapt the relevant themes of the CMW model, we removed those codes that were not reflected in the transcript.

## Results

A total of 25 semi-structured interviews were conducted with health officials who were involved in community midwives’ training and deployment (n=5), midwifery students (n=5), community midwives working in the district (n=5), community midwives trained but not working in their professions (n=5), and community women (n=5). Tables 1 to 4 describe the demographic details of the study participants.

**Table 1.**
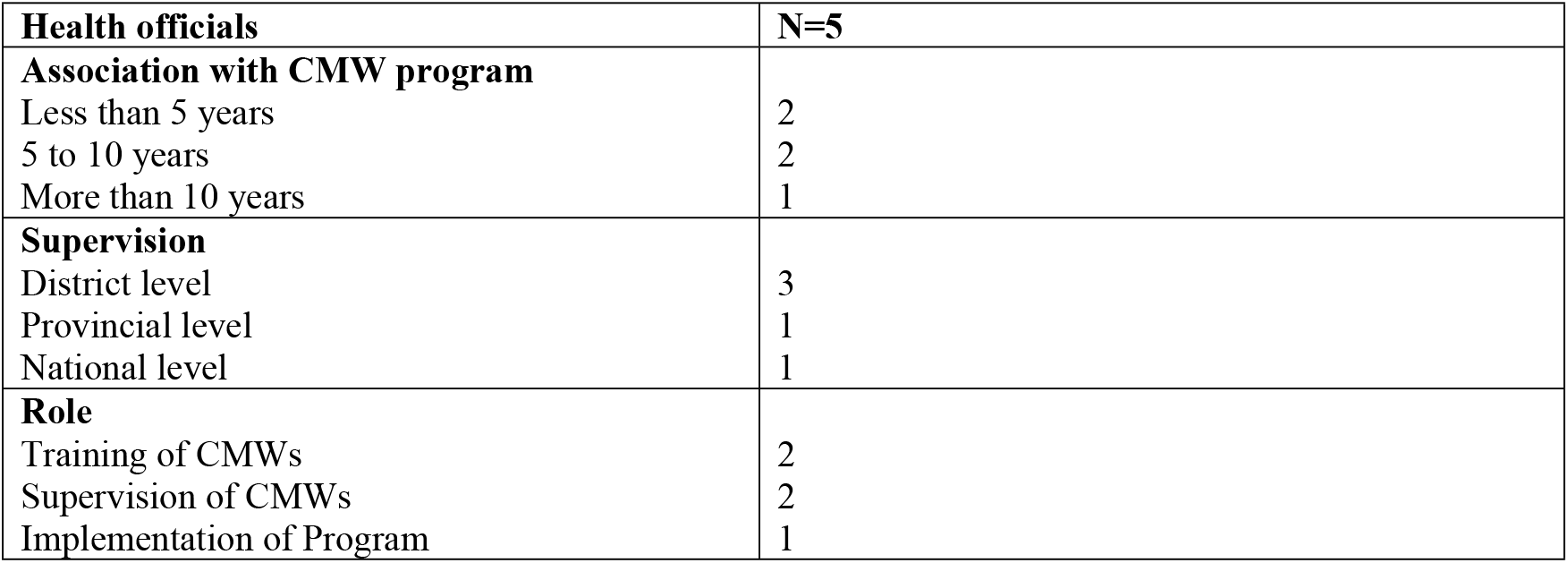
Characteristics of Health Officials.

**Table 2.** Demographic details of midwifery students.

| Midwifery students | N=5 |
| --- | --- |
| **Age** |  |
| 16-25 | 4 |
| 26-35 | 1 |
| **Previous Education** |  |
| Intermediate | 5 |
| **Marital status** |  |
| Unmarried | 4 |
| Married | 1 |

**Table 3.** Demographic details of working and non-working groups of midwives.

| **Midwives** | **N=10** |
| --- | --- |
| **Age** |  |
| 21-25 | 2 |
| 26-30 | 1 |
| 31-35 | 4 |
| 36-40 | 3 |
| **Education** |  |
| High school | 5 |
| Undergraduate | 2 |
| Graduate | 3 |
| **Midwifery course** |  |
| 12 months diploma | 3 |
| 18 months diploma | 7 |
| **Workplace:** |  |
| Non-Governmental Organizations | 4 |
| Private organizations | 5 |
| Health facilities | 1 |
| **Working experience as midwife** |  |
| Less than 5 years | 2 |
| Less than 10 years | 5 |
| More than 10 years | 2 |
| **Marital status** |  |
| Unmarried | 2 |
| Married | 8 |

**Table 4.** Demographic details of community women.

| **Community-women** | **N=5** |
| --- | --- |
| **Age** |  |
| 16-30 | 1 |
| 31-45 | 3 |
| >45 | 1 |
| **Education** |  |
| None | 2 |
| Primary | 1 |
| College | 2 |
| **Antenatal visits for recent pregnancy** |  |
| Less than 3 | 3 |
| 3 to 8 | 2 |
| **Facility approach during recent childbirth** |  |
| None (home birth) | 1 |
| CMWs birthing units | 1 |
| BHU, RHC (other primary facilities) | 1 |
| Secondary and tertiary care hospitals | 2 |

All the study participants were contacted by the research team using a purposive sampling approach and they provided agreement to participate before conducting interviews with them. Based on the data collection and thematic analyses involving the four domains of the CMW model, two overarching themes were identified: (I) community acceptance and support, and (II) dynamics between CMWs and health care providers. The themes and subthemes, and illustrative quotes are presented in Table 5.

**Table 5:**
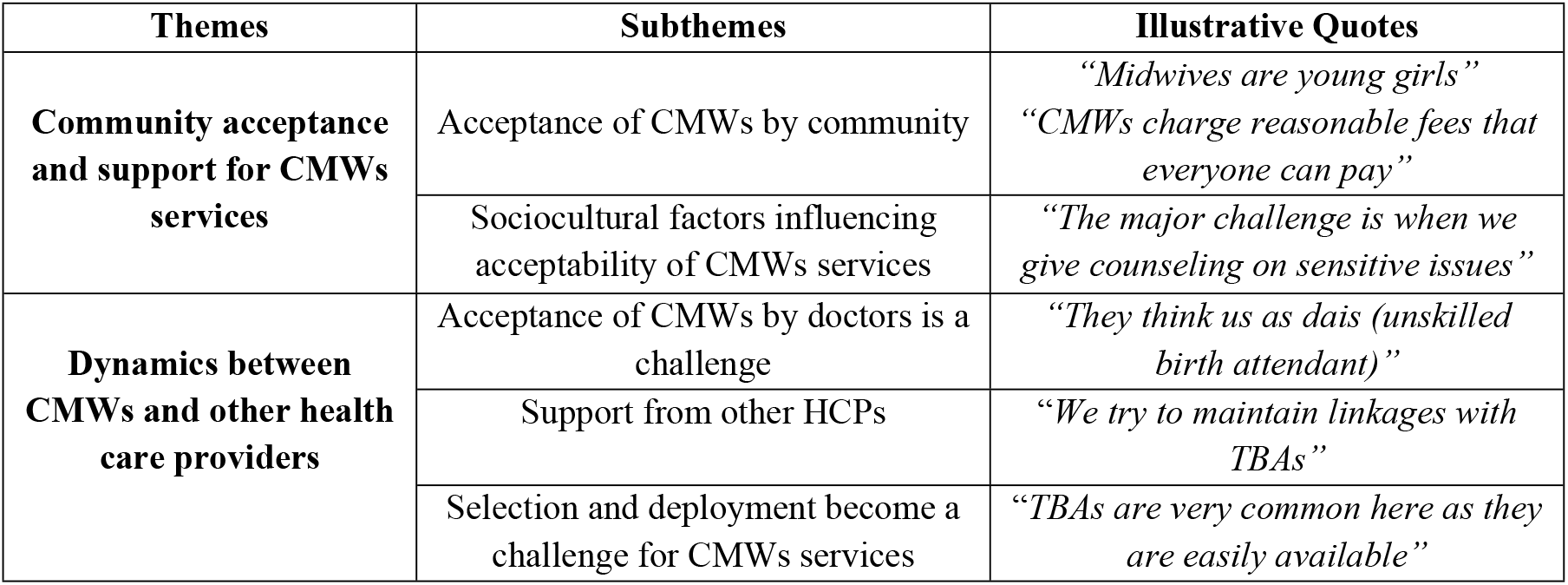
Themes and subthemes.

### Community acceptance and support for CMWs services

#### Acceptance of CMWs by community

A majority of midwifery students who were interviewed were unmarried and were under the age of 25 years. The eligibility criterion to enroll in the midwifery program requires the completion of 12 years of schooling. Most of the girls get enrolled in the program at the ages of 18 to 20 years and are certified to provide services after the successful completion of two years. The novice CMWs being young and unmarried were considered naive by community women to deal with reproductive health matters, which is considered as something related to being married or to an older age group.

> *“We try to enroll young and competent girls who show up their commitment to work in communities (KII-principal, midwifery school)*
>
> *“Midwives are young girls. They are not competent for services. They go and study, but they don’t have practical skills to perform deliveries” (IDI-Community woman)*
>
> *“The young age of community midwives is not acceptable by communities. They think that unmarried girls and young girls do not have any experience to deal with these matters. Of course, experience is always given importance in comparison to the education in any of the fields” (KII-District Health Officer)*

On the other hand, a community woman acknowledged CMWs’ diverse role in the provision of maternal and newborn services that make them significant as they support women throughout their pregnancy and childbirth process.

> *“CMWs educate patients, listen to patient concerns, talk to their families, help them with other services such as ultrasound and manage them according to the problem” (IDI-community woman)*

Besides, the community’s affordability for CMWs services was identified as a major facilitator for their services. As CMWs are part of their own communities where they work, they charge fees for services according to individual’s affordability. Most of the community women mentioned that CMWs charge less fees compared to doctors.

> *“CMWs charge reasonable fees that everyone can pay. It is 5 to 10 times less than the doctor’s fees” (IDI-Community woman)*
>
> *“People do not want to go to doctors because doctors do not provide them adequate time to listen to their concerns and they charge a lot for their services” (KII-Director, general nursing, and midwifery)*

However, a few CMWs reflected that they face challenges in convincing women to refer them to a better-equipped health facility in cases of high-risk pregnancies or when suffered from pregnancy complications. One CMW mentioned:

> *“It is very difficult to explain to them about the referral. They say that if we had to go there why we have visited you and what are you here for” (IDI-Working midwife)*

Another reason for refusing referral according to midwives’ viewpoints is the lack of privacy in labor rooms of hospitals as cited by a CMW:

> *“In our area, there is one big hospital that is civil hospital Thatta where multiple patients deliver in a single labor room. Most women do not prefer to go to hospitals because of that. We conduct delivery in a single private room that’s why people prefer to come to midwives than going to the hospital” (IDI-working midwife)*

Contradictory to this, community women showed up their interest in having childbirth at home instead of covering the distance to reach any health facility such as a midwife’s birthing station. They voiced their concerns regarding the non-availability of transportation and increased costs to move towards other facilities. They preferred traditional birth attendants as they were easily available and provided home services at low costs.

> *“Dais (TBAs) in our areas are easily approachable and we don’t have to pay anything to reach any other facility. They charge very less in comparison to anyone else” (IDI-community women)*
>
> *“To increase skilled birth attendance, we started training TBAs to link them with CMWs but this strategy didn’t work because people are reluctant to go out for deliveries. TBAs provide home services and even if CMWs would provide them home services, they would prefer a TBA over CMW because of low cost and experience in childbirth process” (KII-Coordinator, MNCH program)*

When exploring community women’s preferences, many women (3/5) were unaware of services provided by community midwives, and they informed that there were no CMWs birthing stations established in their areas.

> *“I have no idea about if we have any birthing station in our area, we have some small setups of clinics and LHVs run those clinics but haven’t heard about any community midwife who is working nearby” (IDI-community woman)*

#### Sociocultural factors influencing acceptability of CMWs services

Many social and cultural issues impede the acceptability of CMWs in rural areas. Most of the CMWs recognized sociocultural issues as a hindering factor for CMWs practices. The non-acceptance of CMWs families to establishing birthing stations at homes, the family’s restrictions to allow CMWs to work in hospitals because of female mobility issues, and the reservations of CMWs to allowing males (husbands of pregnant women) at their birthing stations were some of the identified impeding factors for CMWs services.

> *“The people in our village and our family don’t allow us to work in a male dominated area because of security issues and our cultural norms” (IDI-Non-working CMW)*

Moreover, CMWs’ counseling and educational services over sensitive issues like family planning, pregnancy termination, or abortion were disregarded by the community people.

> *“Working as a midwife is a challenge itself, acceptance among the community is the major challenge and this is when we give counseling on family planning or abortion. It’s because we’ve taboos associated with these topics” (IDI-Working midwife)*
>
> *“As midwives provide services to their local communities, they have to face many cultural challenges” (KII-general secretary-MAP)*

### Dynamics between CMWs and other health care providers

#### Acceptance of CMWs by doctors is a challenge

Most of the respondents acknowledged that CMWs strive to maintain a positive relationship with every health care provider working in the community and especially with the doctors who provide pregnancy and delivery care. However, their efforts are not acknowledged by the doctors, and they consider CMWs as ‘*unskilled birth attendants (Dais)’*, and not trained healthcare providers. Doctors also misinform community people about their services as acknowledged:

> *“Women Medical Officers misguide people against CMWs if they refer any complicated case to doctors. The referred woman when is delivered by the doctor, the whole credit is taken by the doctor, and they provoke patients about the complications. When the same patient goes back to her home, she gives that message to everyone in the community that CMWs are uneducated and non-qualified to conduct deliveries” (KII-Coordinator MNCH program)*
>
> *“At their level, we are nothing. They think of us as dais (unskilled birth attendants). Some of them support us and they think that there is someone who has the knowledge, but some say to their clients to visit them and not us as they think we are dais and they think that we have not learned much” (IDI-Working midwife)*

#### Support from other HCPs

CMWs stated that they try to build linkages with health facilities and other health care providers working in the district to establish their work, for example with sonologists and laboratories to refer their clients for essential services. Some of the CMWs also reported that they maintain good relations with TBAs, and they provide a monetary share to TBAs from service charges for delivery and childbirth, if any client is referred to them by TBAs.

> *“We have good linkages with Sonographers and laboratories to referring patients for services” (IDI-Working midwife)*
>
> *“We try to maintain linkages with TBAs. We provide them some share if they refer any patient to them for delivery” (IDI-Working midwife)*

Lady Health Workers (LHWs) play an essential role in providing health services to communities, especially in rural areas to those people who have difficulty in access to health services. Every CMW in district Thatta is consigned to five Lady Health Workers (LHWs) of the specific area.

LHWs indicate and register individuals and households in the area and refer clients to nearby CMWs for ANC, PNC, delivery care, and other maternal services to increase skilled birth attendance in rural areas. However, the coordination between LHWs and CMWs continued to be discordant. This is because LHWs are ineligible and non-licensed to provide maternal and newborn services and the professional rivalry creates challenges for CMWs to coordinate with LHWs.

> *“Under one CMW, there are 5 LHWs to support each other. The registration of pregnant women is carried out by the LHWs. They register each person in the village, the children, the adults, and everyone. The bad luck is that we couldn’t strengthen the coordination between CMWs and LHWs” (KII-district health coordinator)*

#### Selection and deployment become a challenge for CMWs services

Thatta is divided into urban and rural geographical zones. The urban zone of Thatta has an increased number of doctors, midwives, and health care providers [18]. The increasing availability and accessibility to doctors and already working CMWs in urban regions challenges newly deployed CMWs in establishing their birthing stations.

> *“I live in a town where every second person has done midwifery and MBBS. They are available in every street. We cannot establish a good setup here because there are a lot of people in this profession” (IDI-Non-working CMW)*
>
> *“We have an admission committee to enroll girls to CMWs from those areas where there is the unavailability of birthing stations, but we still need to strengthen this as we see that there are many gaps (KII-principal, midwifery school)*

CMWs’ deployment in areas with increased dispersion of TBAs’ is a barrier to their services. Most of the community women preferred TBAs for delivery care over CMWS as they believe that young, unmarried, and inexperienced CMWs cannot provide efficient maternal services compared to an experienced TBA who provides low-cost services.

> *“TBAs are very common here as they are easily available, and they are very experienced to conduct deliveries in our village” (IDI-Community woman)*
>
> *“The strategies to increase skill birth attendance through CMWs is still lacking as people still prefer TBAs over CMWs” (KII-coordinator, MNCH program)*

## Discussion

CMW program is an important part of the MNCH program, and it can improve skilled care birth attendance in rural areas of Pakistan if accepted by communities and other health care providers. This study identified facilitators and barriers to community midwives’ acceptance in the context of district Thatta.

The young age of a midwife to establish her own birthing station is not acceptable by the communities. The maternal and neonatal services provided by young and unmarried girls is a hindering factor to gain the trust of the community, as they perceive them to be inexperienced and uninformed about reproductive health matters. The enrollment of girls in the CMW program and becoming eligible to provide maternal and neonatal services as CMW at a very young age is not accepted by communities. Most of the CMWs in the district are unmarried and community women consider them novice to assist them with childbirth processes. Similar findings on the lack of community trust for CMWs services have also been identified in a qualitative study carried out on CMWs working in different parts of the province of Sindh [24]. Besides, these findings have been also reported from regions outside Pakistan such as Nigeria and Niger reported similar results on the non-acceptance of midwifery personnel due to their typically young age [25-26].

However, the CMWs cost for services is acknowledged by community people as they charge two to three times less fees than medical doctors and obstetricians. Moreover, CMWs are better informed about the background and financial state of their local communities, and they consider the cost of services accordingly. This is one of the significant aspects that attracts community women to utilize CMWs services. The relatively reduced cost of services of CMWs in comparison to doctors has been reported as a key facilitator in other LMICs including Africa[27]. Some experts have also noted that high-income countries with the lowest intervention rates, best outcomes, and lowest costs have integrated midwifery-led care into their health care systems [28]. Evidence from high-income countries found such models to be a cost-efficient way to improve health outcomes, reduce medical interventions and increase satisfaction with care [29].

Community midwives provide diverse roles such as patient assessment, health education, advocacy, and referral of high-risk pregnancies. The varied role of CMWs fascinates community women to utilize their services however, the awareness of CMWs’ services to their communities is a problem. The high accessibility and readiness of traditional birth attendants to perform deliveries in the rural communities hinder access of women to CMWs and they prefer to deliver babies at *‘home’* rather than reaching out to any facility or birthing stations. The approach to TBAs during pregnancy and childbirth is found in most rural areas in all provinces of Pakistan [30]. While rates of home delivery have decreased in LMICs, about 25% of deliveries worldwide still occur outside a health facility [31]. TBAs still continue to play significant roles in maternal health care in rural and deprived communities where there is a shortage of skilled birth attendants. The findings from Ghana informed that women traditionally prefer to deliver at home because it is cheaper and easier as women who deliver at home receive social support from their extended families and do not have to pay much for the delivery services [32].

The social and cultural system is another factor that hinders the acceptability of CMWs in rural areas. CMWs find challenges to educate community people about important yet sensitive maternal health matters such as family planning and abortions, which are not given importance, especially in rural areas, and addressing these become a challenge for community midwives.

Moreover, male family members don’t allow females to work due to the patriarchal system followed in rural areas of Pakistan. CMWs are not allowed to establish birthing stations at their homes and are not allowed to work in community clinics or maternity settings due to female mobility issues which are the cause of underutilization of midwifery services in rural areas of Pakistan. The female mobility issues in Pakistan have underestimated female empowerment and this hinders female development in every aspect such as social, economic, and career development [33].

Besides, the community’s and the healthcare providers’ support for CMWs’ services is insignificant in terms of the acceptability of services. CMWs struggle to gain their status in communities where TBAs and doctors are recognized to deliver maternal and newborn services. The communication and coordination between CMWs and doctors are negligible. Doctors consider them unskilled professionals and they convey this perception to communities which becomes a cause of their non-acceptability by the community. The conflict between CMWs and doctors for utilization of maternal and newborn services has been identified in different parts of the world such as in a study conducted in Australian public hospitals [34]. Despite much progress in the legitimization of the midwifery profession in many high-income countries, these countries also face disagreements among maternity care professionals regarding midwifery autonomy and scope of their practice [35]. Maternity care and services require close cooperation between obstetricians, midwives, physicians, and nurses [36].

The functioning of CMWs in the district depends on coordination with LHWs for the referral of pregnant women to CMWs. However, the lack of coordination between LHWs and CMWs impedes their services. This lack of coordination between two cadres is presumed due to the professional rivalry between the two cadres. Yet, professional jealousy is a typical phenomenon in healthcare and has been experienced in every health setting from a smaller setting to a larger setup [37].

It has been identified that the district Thatta is divided into rural and urban zones and most of the girls were enrolled from the urban regions where there was already a high saturation of healthcare providers such as medical doctors, CMWs, and LHVs. The rural regions of the district still suffer from issues of non-availability of birthing stations and even if CMWs were available in those regions, they were not functional due to the increased approach of the community towards TBAs for deliveries. This is due to a lack of planning of program implementers to execute this significant program in rural communities for the purpose the program was evolved. A similar finding has been reported in a study carried out on capturing the views of MNCH program specialists who provided their insights on the malfunctioning of various vertical programs after the devolution of the federal government to provincial governments [38]. Ideally, devolution provides increased opportunities for provinces to introduce and prioritize health policies. Countries like Indonesia and Kenya provide useful examples of improved health systems following devolution [39-40]. The health services delivery and health governance in these countries have improved after the devolution [39-40]. Devolution has transformed the power and decision-making authorities at the subnational level and has provided increased opportunities to prioritize preventive health services rather than curative health services [41].

However, the reform in Pakistan faced a number of challenges due to a lack of defined national policies and unorganized provincial health strategies [38].

Our study has several strengths. First, the exploratory design of this study helped us to understand the factors that influence CMWs’ acceptability from all levels of the CMW program from organization to implementation. Second, the rigor of the study was achieved by enhancing the credibility of the findings from different data sources including health officials, midwives, students, and community women.

The study also has some limitations. First, the data collection was done through telephonic interviews because of the pandemic situation and the facial expressions and physical observations were therefore missed during the data collection. This also limited us to capture data from the participants who had no access to mobile phones. Secondly, the study focused only on the perspective of the participants who are a direct or indirect part of the MNCH or CMW program. The interviews with doctors, LHWs, and TBAs of the rural areas would have provided more insights and understanding of the current processes to strengthen the findings.

In order to address the effective utilization of CMWs services, the study recommends strengthening the program implementation at all levels. Firstly, the study provides insights to program implementers to rearrange the selection and deployment of CMWs. They should be selected from the areas where there is a need for CMWs and not from the areas with a saturation of doctors, working midwives, and TBAs. Secondly, the placement of CMWs must be linked with other MNCH activities to facilitate referral, promote health education, and create awareness of this significant cadre among health care providers and in the communities. Thirdly, the recognition of CMWs as professionals and skilled workers should be emphasized through different platforms to highlight their important role in maternal and neonatal services provision. Lastly, efforts to increase female empowerment are required to sustain services provided by skilled care workers such as CMWs.

## Conclusion

In conclusion, community midwives can play a vital role in reducing maternal and newborn mortality rates and in achieving progress in health service delivery. However, the findings suggest that these improvements cannot be made until this cadre will be accepted by community people and other health care providers. The study provided an extensive understanding of the factors influencing CMWs acceptability in district Thatta.

The insights to program implementers to rearrange age criteria for selection and deployment of CMW and the program implementers need to work on strategies and interventions to develop the community’s and health care providers’ trust for community midwives’ services. This can help to achieve the aim of introducing the CMW program in Pakistan. However, disregarding community acceptance and absorption of CMWs in the health system, the aim of the development of this cadre cannot be achieved. The CMW program has been engrossed in the health system of Pakistan but has not yet fully accepted to carry out its roles.

## Data Availability

The datasets were collected and analyzed and can be made available from the corresponding author on reasonable request.

## Acknowledgment

We would like to acknowledge Ms. Arusa Lakhani (School of Nursing, Aga Khan University), Mehmooda Afroz Ali Khowaja (School of Midwifery, Lady Dufferin), Dr. Raheela (School of Midwifery, Thatta) for their assistance in data collection and providing contacts with key stakeholders.

## Ethics

Ethical approval to conduct the study was given by the Aga Khan University Ethical Review Committee (2020-3391-11138). Participants provided verbal consent to indicate their willingness to participate. Voluntary participation and the right to ask any questions and to decline participation at any time were emphasized during the data collection.

## Author Contributions

BK is a student of the MSc Health Policy and Management Programme in the Aga Khan University, Community Health Sciences Department, Karachi, Pakistan. The manuscript has been prepared from her thesis work. The thesis has been supervised by SS. The thesis committee members AF & FA supported the conduct of this research. This qualitative study was conceptualized by BK and SS. BK conducted the study and prepared the first draft of the manuscript. SS, AF, FA & MR provided facilitation during the data collection. SS and AF provided guidance for developing the conceptual framework of the study. SS, AF, FA, QK, and MR reviewed the manuscript several times and provided feedback. All authors have reviewed and approved the final version of the manuscript

## References

1. World Health Organization. Making every baby count: audit and review of stillbirths and neonatal deaths. www.who.int/maternalnewbornadolescent/documents/stillbirth-neonatal-death-review/en/. Geneva: World Health Organization (WHO), 2016.

2. Bauserman M, Thorsten VR, Nolen TL, Patterson J, Lokangaka A, Tshefu A, Patel AB, Hibberd PL, Garces AL, Figueroa L, Krebs NF. Maternal mortality in six low and lower-middle income countries from 2010 to 2018: risk factors and trends. Reproductive health. 2020 Dec;17(3):1–0.

3. Geller SE, Koch AR, Garland CE, MacDonald EJ, Storey F, Lawton B. A global view of severe maternal morbidity: moving beyond maternal mortality. Reproductive health. 2018 Jun;15(1):31–43.

4. World Health Organization. Maternal mortality: evidence brief. World Health Organization; 2019.

5. Moyer JD, Hedden S. Are we on the right path to achieve the sustainable development goals?. World Development. 2020 Mar 1;127:104749.

6. Yaya S, Ghose B. Global inequality in maternal health care service utilization: implications for sustainable development goals. Health Equity. 2019 Apr 26;3(1):145–54.

7. World Health Organization. Definition of skilled health personnel providing care during childbirth: the 2018 joint statement by WHO, UNFPA, UNICEF, ICM, ICN, FIGO and IPA. World Health Organization; 2018.

8. Edmonds JK, Ivanof J, Kafulafula U. Midwife led units: transforming maternity care globally. Annals of global health. 2020;86(1).

9. Murrary-Davis B, Hutton EK, Carty E, Kaufman K, Butler M, Martin G, Lam A, Bi M, Belanger J, Campbell A, Robertson A. Comprehensive Midwifery-The role of the midwife in health care practice, education, and research: An Interactive Guide to the Theory and Evidence of Practice.

10. Berg M, Ólafsdóttir ÓA, Lundgren I. A midwifery model of woman-centred newbornbirth care–In Swedish and Icelandic settings. Sexual & Reproductive Healthcare. 2012 Jun 1;3(2):79–87.

11. Kachikis A, Moller AB, Allen T, Say L, Chou D. Equity and intrapartum care by skilled birth attendant globally: protocol for a systematic review. BMJ open. 2018 May 1;8(5):e019922.

12. National Maternal NaCHP. Program PC-1. Islamabad: Maternal, Neonatal and Child Health Program, MInistry of Health, Government of Pakistan; 2007

13. Majrooh MA, Hasnain S, Akram J, Siddiqui A, Memon ZA. Coverage and quality of antenatal care provided at primary health care facilities in the ‘Punjab’province of ‘Pakistan’. Plos one. 2014 Nov 19;9(11):e113390.

14. Shahid S. Midwives in low-resource settings. British Journal of Midwifery. 2020 Nov 2;28(11):796–8.

15. Jan R, Lakhani A, Rattani S, Lalji L, Mubeen K, Jaffer MQ. Can CMWs sustain quality services and high coverage as private providers in Chitral? A three-year prospective qualitative study. Journal of Asian Midwives (JAM). 2019;6(2):23–39.

16. Khowaja BM, Feroz AS, Saleem S. Factors influencing utilisation of services provided by community midwives and their non-retention in district Thatta, Pakistan: a qualitative study protocol. BMJ open. 2022 Jul 1;12(7):e052323.

17. Khowaja BM, Feroz AS, Saleem S. Facilitators and barriers influencing utilization of services provided by community midwives in district Thatta, Pakistan: a qualitative exploratory study. BMC pregnancy and childbirth. 2022 Dec;22(1):1–2.

18. http://rdpi.org.pk/wp-content/uploads/2016/06/District-Profile-Thatta.pdf. (2010).

19. National Institute of Population Studies (Pakistan), Macro International. Institute for Resource Development. Demographic, Health Surveys. Pakistan demographic and health survey. National Institute of Population Studies; 2012.

20. Sahito NG, Channa AA. Bureau of statistics planning and development department of Sindh. Health Profile of Sindh; 2016 Jun 13.

21. Recognized Institutes Of PNC [online], 2022. Available: https://www.pnc.org.pk/PNC_Recognized_Institutes.htm

22. Hennink MM, Kaiser BN. Saturation in qualitative research. Thousand Oaks, CA: Sage Publications Limited; 2020.

23. Belur J, Tompson L, Thornton A, Simon M. Interrater reliability in systematic review methodology: exploring variation in coder decision-making. Sociological methods & research. 2021 May;50(2):837–65.

24. Noh JW, Kim YM, Lee LJ, Akram N, Shahid F, Kwon YD, Stekelenburg J. Factors associated with the use of antenatal care in Sindh province, Pakistan: A population-based study. PloS one. 2019 Apr 3;14(4):e0213987.

25. Wahlström R, Vahidi R, Nikniaz A, Marions L, Johansson A. Barriers to high-quality primary reproductive health services in an urban area of Iran: views of public health providers. Midwifery. 2009 Dec 1;25(6):721–30.

26. Abimbola S, Okoli U, Olubajo O, Abdullahi MJ, Pate MA. The midwives service scheme in Nigeria. PLoS medicine. 2012 May 1;9(5):e1001211.

27. Crisp N, Brownie S, Refsum C. Nursing & Midwifery: The key to the rapid and cost effective expansion of high quality universal healthcare.

28. Tikkanen R, Gunja MZ, FitzGerald M, Zephyrin L. Maternal mortality and maternity care in the United States compared to 10 other developed countries. The Commonwealth Fund. 2020 Nov 18;10.

29. Choudhary S, Jelly P, Mahala P. Models of maternity care: a continuity of midwifery care. International Journal of Reproduction, Contraception, Obstetrics and Gynecology. 2020 Jun 1;9(6):2667.

30. Omer S, Zakar R, Zakar MZ, Fischer F. The influence of social and cultural practices on maternal mortality: a qualitative study from South Punjab, Pakistan. Reproductive health. 2021 Dec;18(1):1–2.

31. Garces A, McClure EM, Espinoza L, Saleem S, Figueroa L, Bucher S, Goldenberg RL. Traditional birth attendants and birth outcomes in low-middle income countries: A review. InSeminars in perinatology 2019 Aug 1 (Vol. 43, No. 5, pp. 247–251). WB Saunders.

32. Adatara P, Afaya A, Baku EA, Salia SM, Asempah A. Perspective of traditional birth attendants on their experiences and roles in maternal health care in rural areas of northern ghana. International journal of reproductive medicine. 2018 Oct 1;2018.

33. Adeel M, Yeh AG. Gendered immobility: influence of social roles and local context on mobility decisions in Pakistan. Transportation planning and technology. 2018 Aug 18;41(6):660–78.

34. Bradfield Z, Kelly M, Hauck Y, Duggan R. Midwives ‘with woman’in the private obstetric model: Where divergent philosophies meet. Women and Birth. 2019 Apr 1;32(2):157–67.

35. Downe S, Finlayson K, Fleming A. Creating a collaborative culture in maternity care. Journal of midwifery & women’s health. 2010 May 1;55(3):250–4.

36. Reeves S, Pelone F, Harrison R, Goldman J, Zwarenstein M. Interprofessional collaboration to improve professional practice and healthcare outcomes. Cochrane Database of Systematic Reviews. 2017(6).

37. Mohammed EN. Knowledge, causes, and experience of inter-professional conflict and rivalry among healthcare professionals in Nigeria. BMC Health Services Research. 2022 Dec;22(1):1–9.

38. Sarfraz M, Hamid S. Exploring managers’ perspectives on MNCH program in Pakistan: a qualitative study. Plos one. 2016 Jan 12;11(1):e0146665.

39. Cannon BJ, Ali JH. Devolution in Kenya four years on: A review of implementation and effects in mandera county. African Conflict and Peacebuilding Review. 2018 Apr 1;8(1):1–28.

40. Maharani A. Decentralisation, performance of health providers and health outcomes in Indonesia. The University of Manchester (United Kingdom); 2015.

41. McCollum R, Limato R, Otiso L, Theobald S, Taegtmeyer M. Health system governance following devolution: comparing experiences of decentralisation in Kenya and Indonesia. BMJ global health. 2018 Sep 1;3(5):e000939.

